# Data Resource Profile: Genomic Data in Multiple British Birth Cohorts (1946-2001)—Health, Social, and Environmental Data from Birth to Old Age

**DOI:** 10.1101/2024.11.06.24316761

**Authors:** Gemma Shireby, Tim T Morris, Andrew Wong, Nish Chaturvedi, George B Ploubidis, Emla Fitzsimmons, Alissa Goodman, Adelaida Sanchez-Galvez, Neil M Davies, Liam Wright, David Bann

## Abstract

Birth cohort studies have a rich history of contributing to science across disciplinary fields, notably health and social sciences. Here, we introduce a curated resource comprising genomic data from five British birth cohort studies—longitudinal studies with extensive data collected prospectively across life, each deliberately sampled to be nationally representative (born 1946–2001). These contain health and social data from birth to older age, enabling longitudinal and cross-cohort genetically informed research. The Millennium Cohort Study additionally includes data on parents and offspring, enabling within-family analyses. Across five cohorts born in 1946, 1958, 1970, 1989–90, and 2000–2002, 27,432 participants have harmonized, imputed, and quality-controlled genetic data from genotyping arrays covering 6.7 million common SNPs. The Millennium Cohort Study contains over 6,000 mother-offspring pairs and over 3,000 mother-father-offspring trios. Pseudonymized data are freely available to the global research community upon approval of a data access request (https://cls.ucl.ac.uk/data-access-training).

## Data resource basics

### Background: the value of genomic data in multiple birth cohorts

Birth cohort studies have a rich history of contributing to science within and between disciplinary fields, notably the health and social sciences.^1-4^ Here, we introduce a curated resource comprising genomic data from multiple British birth cohort studies—longitudinal studies with rich data collected prospectively across life, each deliberately sampled to be nationally representative (Figure 1). We also outline our platform and steps to aid in cross-cohort harmonised analysis.

**Figure 1.**
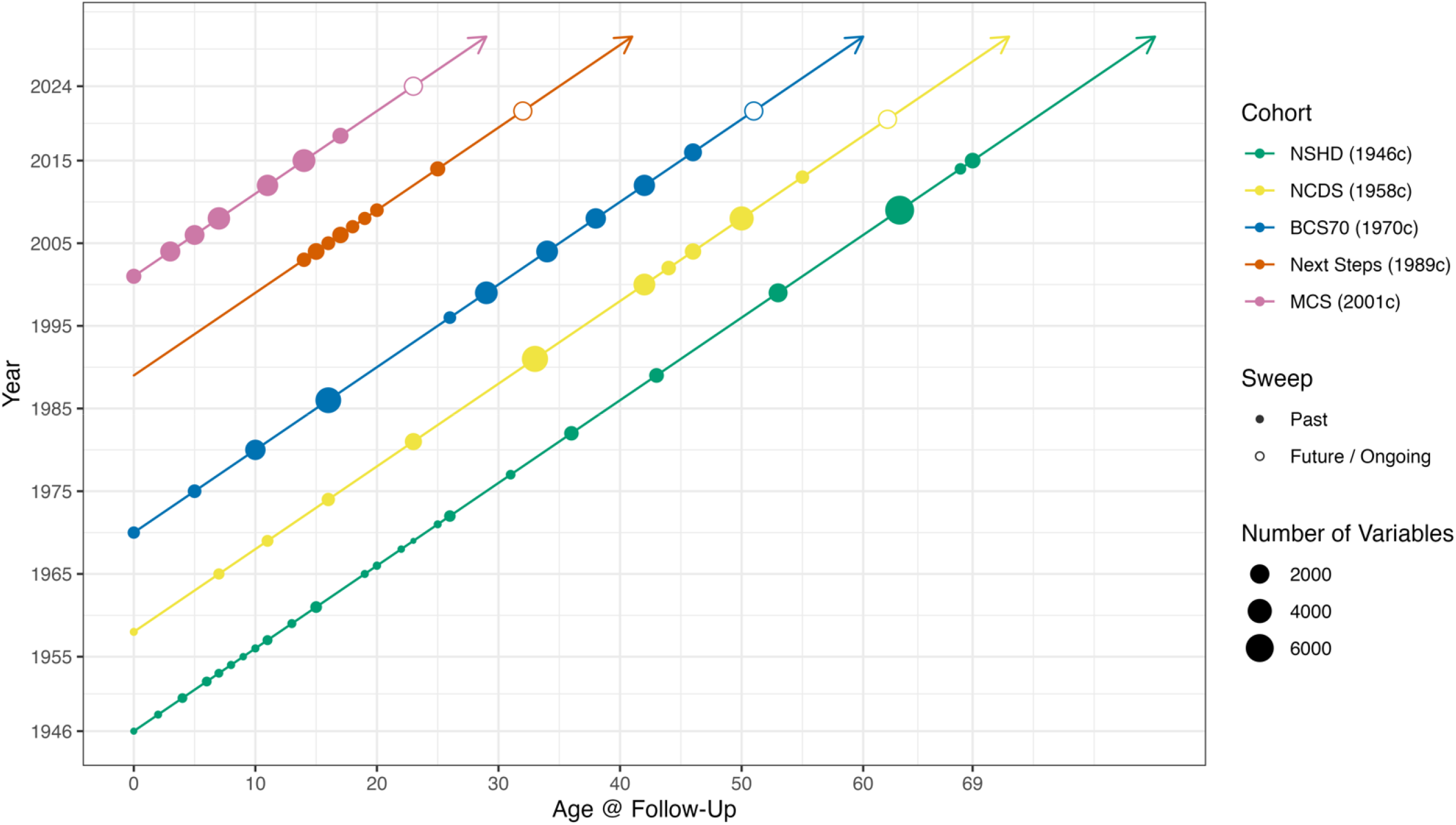
Genomic Data in Five British Birth Cohorts. Note: the number of variables is indicative rather than comprehensive: since values cannot be compared across cohorts due to differences in variable derivation, they indicate the relative availability of variables across age within each cohort.

This paper provides information on the genotyping, imputation, and derived genetic data (polygenic indices (PGIs), also known as polygenic scores) for four national birth cohort studies initiated in 1946 (National Survey of Health and Development, 1946c),^5^ 1958 (National Child Development Study, 1958c),^6^ 1970 (British Cohort Study, 1970c),^7 8^ and the millennium (Millennium Cohort Study, 2001c).^9^ In addition, we include a cohort born in 1989-90 followed up from adolescence (Next Steps, 1989c). Genotyping was conducted for the main cohort participants in each study, enabling population-level longitudinal genetic analyses covering up to 78 years after birth. Furthermore, in the case of the 2001c, the cohort participants’ co-resident (biological) mothers and fathers were also genotyped, enabling powerful family-based genomic analyses.^10 11^

Phenotype data for these cohorts has been described extensively elsewhere; see^5-7 9^ and https://cls.ucl.ac.uk/cls-studies. Briefly, these studies have collected data on a wide range of domains, including physical and mental health, health behaviours (including, in a subset of cohorts, biomarkers and accelerometer data), psychosocial wellbeing, education, employment, cognition, personality, beliefs and attitudes, partnership formation and dissolution, and fertility across the life course. The studies have added further value through extensive data linkage to administrative and other data covering health, schooling, and local area factors (e.g., neighbourhood deprivation). Phenotypic data has been measured regularly across the cohorts (**Figure 1**) from birth through adulthood and into older age (1946c and 1958c). Each is a ‘live’ cohort, with future data collections planned.

## Data Collected

### Sample collection

Saliva samples were used for DNA collection in 2001c (14y) and 1989c (32y); in all other cohorts, blood samples were used, collected at 53y (1946c), 44y (1958c), and 46y (1970c), respectively. Informed consent was obtained from participants and, for 2001c, their parents. Details of the data collection protocols, responses, predictors of response in 2001c has been previously described.^12^

### Genotype data pre-processing

Genotype calling was performed using GenomeStudio (v2.0, Illumina) and quality control was completed using PLINK^13^ 1.9 and 2.0. Samples were read into GenomeStudio (0-1.27% samples excluded) and mapped to a manifest file. Individuals were excluded if they had (i) >2% missing data (1.88%-3.50% samples excluded), (ii) their genotype predicted sex using X chromosome homozygosity was discordant with their reported sex (excluding females with an F value > 0.2 and males with an F value < 0.8) (0.18%-2.14% samples excluded) (iii) they had excess heterozygosity [> three standard deviations (SD) from the mean] (0.36%-1.07% samples excluded), (iv) For related individuals in 1946c, 1958c, 1970c, and 1989c the King algorithm (king-cutoff 0.0884) was employed to identify and exclude one individual from each pair of closely related individuals (3^rd^ degree or closer) (0.19%-0.59% samples excluded). In 2001c, King was utilised to verify family relationships and rectify instances where parents and children were incorrectly paired. In cases where an individual could not be correctly matched to their family, that specific individual was removed from the dataset (0.12% samples excluded). Duplicate samples were removed, retaining those with the higher genotyping rate.

We identified European samples by (i) merging the genotypes with data from 1000 genomes Phase 3, (ii) linkage disequilibrium pruning the overlapping single nucleotide polymorphisms (SNPs) such that no pair of SNPs within 1000□bp had r2□> □ 0.20 and (iii) using an elastic net model to establish which of the super populations the samples fall into (Africans [AFR], Admixed Americans [AMR], East Asians [EAS], Europeans [EUR] and South Asians [SAS]). Although this method puts each sample into the nearest superpopulation, there are still ancestral outliers. We advise these are removed based on principal components. We retain samples from all ancestry and provide a variable to capture this. Before imputation, SNPs with high levels of missing data (>3%), Hardy-Weinberg equilibrium P<1e-6 or minor allele frequency <1% were excluded.

The genetic data were then recoded as vcf files before uploading to the TOPMed Imputation Server which uses Eagle2 to phase haplotypes, and Minimac4 (https://genome.sph.umich.edu/wiki/Minimac4) with the TOPMed reference panel. The genome build was updated to hg38 using LiftOver, implemented within the TOPMed server. This update applied to all data except for Next Steps, which already used the hg38 build. Imputed genotypes were then filtered with PLINK2.0alpha, excluding SNPs with an R2 INFO score < 0.8 and recoded as binary PLINK format. Proceeding with PLINK1.9, samples with >2% missing values, SNPs with >2 alleles,

>3% missing values, Hardy-Weinberg equilibrium P<1e-6 or a minor allele frequency of <1% were excluded (indels have not been excluded). In 1958c 5 chips were QCd separately and combined after TOPMed imputation, where duplicated samples were removed, retaining those which had a better genotyping rate (40% samples excluded), QC checks were run again on the combined sample, checking for related individuals across chips (0 samples excluded). For more details on samples failed please see Supplementary Table 1.

### Genomic data across the British Birth cohorts

Table 1 displays the sample sizes for those who provided a biological sample which subsequently passed imputation and QC’d procedures, as well as three possible denominators: i) those who responded at the age of genotyping (i.e. provided some valid data such as via survey questionnaire); ii) those theoretically eligible for genotyping (e.g., those who had not died or emigrated), which corresponds to the target population of each cohort; and iii) the total cohort (those who had ever responded); Response rates can be calculated using either denominator—differences between studies may arise due to multiple reasons: differences in study scale (e.g., 1946c is smaller), resourcing of each study, and the context each cohort has operated in (e.g., 1989c was historically an Education-focused study operating from within a UK Government department, potentially adversely affecting willingness to provide DNA samples; secular declines in response rates have also occurred in social surveys). Response rates calculated using participants i) responsive at the age of genotyping (e.g., via survey response) and ii) not known to have died or migrated at age of genotyping as the denominator are as follows: 1946c: (92.1%, 64.8%), 1958c (68.2%, 40%), 1970c (65.2%, 33.8%), 1989c (21.5%, 10.2%), and 2001c (66.1%, 42.2%).

**Table 1.** Genomic Data in British Birth Cohorts.

| Cohort | Age biological sample taken for genotyping (years) | N imputed + QC'd genetic data | N responsive at age of genotype* | N alive / in UK at age of genotyping** | N total birth cohort*** | Genotyping array(s) | Post imputation coverage |
| --- | --- | --- | --- | --- | --- | --- | --- |
| 1946c NSHD | 53<br>(then at 60-64, 69, 78) | 2,794 | 3,035 | 4,313 | 5,362 | Illumina MetaboChip<br>Illumina DrugDev<br>Illumina NeuroX2 | 7,816,646 |
| 1958c NCDS | 44<br>(then at 61-63) | 6,396 | 9,377 | 15,971 | 18,558 | Illumina 1.2m<br>Infinium 550<br>Infinium 550k<br>Affymetrix v6<br>Illumina HumanQuad 660 | 7,545,708 |
| 1970c BCS | 46 | 5,598 | 8,581 | 16,577 | 17,006 | Illumina GSA array (v3) | 8,640,849 |
| 1989-90c Next Steps | 32 | 1,568 | 7,279 | 15,358 | 16,122 | Illumina GSA array (v3) | 8,084,945 |
| 2001c Millennium cohort (members) | 14 | 7,841 | 11,859<sup>‡</sup> | 18,575 | 19,517 | Illumina GSA array (v1) | 8,720,874 |
| 2001c Millennium cohort (mothers) | 28-65<br>(median 44) | 7,781 | 11,574 | <sup>§</sup> | 19,162 | Illumina GSA array (v1) | 8,720,874 |
| 2001c Millennium cohort (fathers) | 29-82<br>(median 47) | 4,635 | 10,414 | <sup>§</sup> | 19,162 | Illumina GSA array (v1) | 8,720,874 |

Note: values may differ in future (e.g., if participants withdraw consent or greater genotyping coverage is obtained). For updated sample sizes, please see: https://cls-genetics.github.io/
\*Those who provided some valid data (e.g., via survey questionnaire)
\*\*Those not known to have died or migrated at the time of biological sampling, including historical prior refusals (In Next Steps also excludes n=22 participants who are in prison, and includes only participants living in England per coverage of this study).
\*\*\*Total birth cohort, inclusive of subsequent additions (e.g., during childhood sweeps). For 2001c, the total for mothers is lower than participants due to multiple births. For BCS, the total birth cohort number stated excludes those living in Northern Ireland who were sampled at birth but not followed up.
<sup>§</sup> Numbers alive at genotyping and in UK are not provided for 2001c parents as they are not the main cohort participants.
Response frequencies for each cohort and sweep can be obtained from the UKDS for the 1958c (SN: 5560), the 1970c (SN: 5641), 1989-90c (SN: 5545), and 2001c (SN's: 4683, 5350, 8156, 8172).

### Within-family genomic data in the 2001c

Genetic data was also obtained from the co-resident (biological) mothers and fathers of 2001c members at the same time as the participants. This collection of family genomic data enables the 2001c to be used for powerful family-based genetically informed analyses (see below). These samples (Table 2) were processed at the same time and in the same way as those for the participants, being genotyped on the same Illumina GSA array (v1) chip: 7,781 study mothers and 4,635 study fathers provided samples that passed imputation and QC. Biological sample provision was nested within families, meaning that many of the participants who provided a sample also had a mother or father who provided a sample, and 40% had data from a complete family trio. This resulted in 6,431 offspring + mother duos, 3,804 offspring + father duos, and 3,119 offspring + mother + father trios.

**Table 2.**
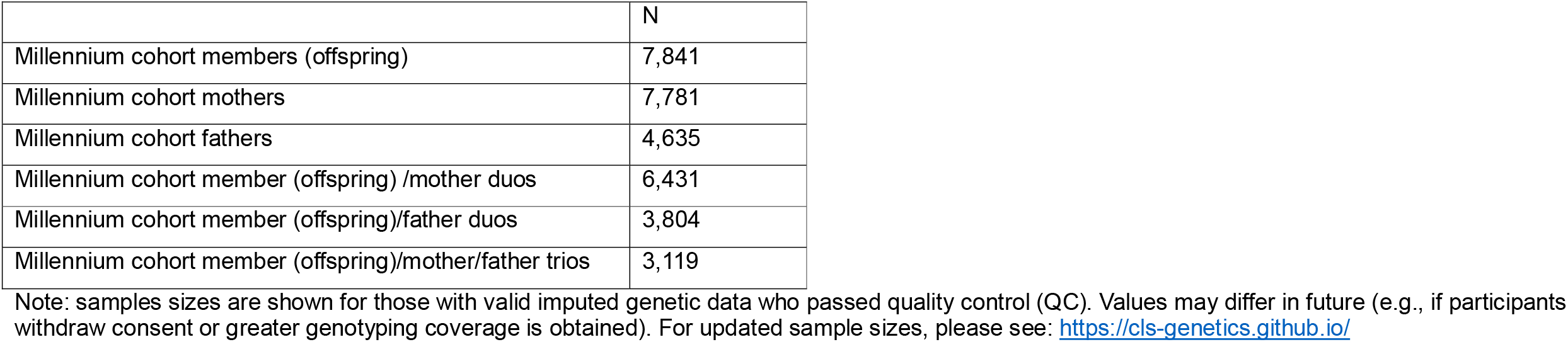
Genomic Family Data in the Millennium cohort study: sample sizes for each familial configuration.

|  | N |
| --- | --- |
| Millennium cohort members (offspring) | 7,841 |
| Millennium cohort mothers | 7,781 |
| Millennium cohort fathers | 4,635 |
| Millennium cohort member (offspring) /mother duos | 6,431 |
| Millennium cohort member (offspring)/father duos | 3,804 |
| Millennium cohort member (offspring)/mother/father trios | 3,119 |

Note: samples sizes are shown for those with valid imputed genetic data who passed quality control (QC). Values may differ in future (e.g., if participants withdraw consent or greater genotyping coverage is obtained). For updated sample sizes, please see: https://cls-genetics.github.io/

### Polygenic index generation

Polygenic indices (PGI) were calculated for various health and social traits (Table 3) which we plan to update periodically in future as more predictive PGIs become available or GWAS on additional traits well suited to cohort research. For an up-to-date list of available PGI please see the CLS genomics page at https://cls-genetics.github.io/docs/PRS.html. We used an additive scoring method summing trait-associated variants in each cohort, weighted by their effect size estimates from GWAS. The number of effect alleles were multiplied by either the log odds ratio or beta coefficient for each SNP. PGI were first generated using PRSice2 (v2.0)^14^ which ‘clumps’ the GWAS summary statistics such that the variant with the lowest p value in each linkage disequilibrium (LD) block is retained. PGI were calculated in each dataset for each individual at two GWAS p-value thresholds: genome-wide significance (p< 5e-08) and suggestive significance (p< 1e-05). The set of significant SNPs included in the genome-wide significant (p < 5e-08) analysis was derived from the genome-wide significant associations reported by the authors in their respective GWAS publications. To identify the list of SNPs at suggestive significance, clumping was used to remove SNPs in LD with each other. We selected SNPs with the lowest p-value within 250kb, removing SNPs with an r^2^ threshold > 0.1 PGI were generated using two methods: first, utilising all available SNPs in each cohort, and second, employing a harmonized set of SNPs (N=6,702,716) common across all cohorts. Before analysis, it is recommended that PGI be standardised to have a mean of 0 and SD of 1, and therefore, the interpretation is in units of SDs.

**Table 3.** Polygenic indices curated in British birth cohorts.

| Domain | Trait |
| --- | --- |
| Physical health / anthropometrics | Addictive behaviour/ substance abuse |
|  | Age at initiation of smoking |
|  | Age at menarche |
|  | Age at menopause |
|  | Asthma |
|  | Birth weight |
|  | Blood pressure |
|  | Body fat percentage |
|  | Body Mass Index |
|  | Body Mass Index (childhood) |
|  | Coronary artery disease |
|  | C-reactive protein measurement |
|  | Fasting blood glucose measurement |
|  | Grip strength measurement |
|  | HbA1c measurement |
|  | Hypertension |
|  | Rheumatoid arthritis |
|  | T1 Diabetes |
|  | T2 Diabetes |
|  | Waist circumference |
| Mental health and cognition | Anxiety |
|  | ADHD |
|  | Alzheimer's Disease |
|  | Autism spectrum disorder |
|  | Bipolar disorder |
|  | Cognition |
|  | Hippocampal volume |
|  | Major depressive disorder |
|  | Parkinson's disease |
|  | Schizophrenia |
| Health/ health behaviours | Alcohol consumption |
|  | Cigarettes per day |
|  | Diet |
|  | Drinks per week |
|  | Smoking |
| Social outcomes | Education |
|  | Household Income |
|  | Human Longevity |
|  | Parental Lifespan |
| Personality | Agreeableness |
|  | Conscientiousness |
|  | Openness to experience |
|  | Neuroticism |
|  | Loneliness |

Note: traits included may increase in future (e.g., as underlying GWAS studies increase in size). For updated information, please see: https://cls-genetics.github.io/

### Data resource uses

In the context of genetically informed research, birth cohorts played key early roles in understanding the genetic underpinning of common diseases (Wellcome Trust Case Control Consortium, established 2005).^15^ In recent years, there has been an explosion of interest in and use of large-scale biobanks drawn from non-representative samples such as the UK Biobank (N=500,000). Since the UK Biobank, other biobanks have been created internationally, and ever larger biobanks are undergoing data collection (e.g., Our Future Health in the UK (target N=5m), All of Us in the US (target N=1m). How can these smaller studies add value in the context of large biobanks and the broader scientific literature?

First, longitudinal cohort studies enable study across the entire lifespan; follow-up is from birth to older ages. This enables the investigation of genetically informed research with a life course framework. For example, do genetic contributions to traits differ across life? Recent work has investigated this with respect to body mass index^16 17^ and blood pressure,^18^ but given the paucity of life course datasets combined with genetics it remains unclear how such patterns differ for other health or social traits, or how gene*environment interactions differ across life. Where power is insufficient in a given cohort, cohorts can be pooled.^19^

Second, longitudinal cohort studies are interdisciplinary resources; their rich health and phenotypic data enable testing of many research questions impossible with limited cross-sectional or questionnaire data from a large biobank. This will enable new evidence to be brought to light on traits well studied in cohort studies, for example, using genetic variants as exposures, confounding variables, or as instrumental variables to mitigate reverse causality / confounding.^20^ For those more familiar with larger-scale biobanks, it will enable the study of phenotypic data that are typically unmeasured or measured poorly in medically oriented biobanks. For instance, child development, personality, cognitive ability, socioeconomic measures across life, and well-being. Their prospective measurement avoids issues relating to retrospective recall bias.

Third, the use of multiple longitudinal cohorts enables comparative research across time, enabling the scientific study of generational change.^4 21^ Several phenotypes have dramatically changed across the latter half of the 20^th^ century, including smoking, BMI and education. For many outcomes, genetic variants’ absolute and relative importance may depend on the societal context in which genes are expressed (e.g., as has been suggested for education^22 23^ or BMI^24^). The provision of data that has been imputed and QC’d in a routine manner across the cohorts means that these data can be analysed consistently and efficiently.

Fourth, their sampling design and national representation aid greater generalisability to the target population and understanding of the broader role of selection bias in genetically informed research than is possible in other large-scale biobanks. While attrition remains a concern across longitudinal studies (as does non-response in cross-sectional studies^25^), there is increasing evidence that the rich data obtained throughout early life in cohorts can be used to reduce bias due to attrition in analyses.^26^ Future work could use these datasets to empirically test the importance of selection bias on larger-scale studies.

Finally, the youngest of the cohorts we include contains genetic data on family trios (mothers, fathers, and cohort member offspring). This enables the separation of direct and indirect genetic effects on outcomes through adjustment for parental genotypes^10 11^ and modelling of parental assortment.^27-30^

### Strengths and limitations of the data resource

Strengths include the consistent approach to data quality control and imputation, and features of the data detailed above—including rich data across life, multiple generations, national representative sampling frame, and family-based genomic data (in 2001c).

Limitations of the genomic data resources in the British Birth Cohorts include non-response, which reduces statistical power and may bias analyses. This can be mitigated (with assumptions) using principled approaches to handle missing data such as multiple imputation or analytical weights.^26 31 32^ As in other harmonisation initiatives, information loss typically results when analysing multiple cohorts together using the same SNP coverage—coverage is limited to the lowest common denominator across included cohorts. Given the broadly high coverage across the harmonised datasets, this is likely to have low impact. For example, using a genome-wide significant PGI for educational attainment and observed education at age 33 in 1958c, the incremental r^2^ of a PGI (above sex and principal components) derived from SNPs harmonised across the cohorts is 9.17% compared with 9.21% for PGI derived from unharmonised SNPs. Conducting sensitivity analyses in cohorts without this restriction (i.e., the largest available coverage without any restrictions) may inform us of the potential implications. Finally, the sample sizes of non-European ancestry are small in adult cohorts born before migration into Britain in the later 20^th^ century (i.e. 1946c, 1958c, and 1970c). However, the Millennium and Next Steps cohorts included oversampling of ethnically diverse areas enabling genetically informed analyses across diverse ancestry groups.^12^ Pooling analyses across multiple cohorts may mitigate power issues if some ancestral sub-groups are insufficiently large in a single cohort.

## Data Availability

Pseudonymised data are freely available to the research community upon approval of a data application request.

https://cls.ucl.ac.uk/data-access-training

https://skylark.ucl.ac.uk

## Data resource access

Pseudonymised data are freely available to the research community upon approval of a data application request. Access is governed by the CLS Data Access Committee (CLS DAC). Researchers may also apply to use stored biological (e.g., blood, saliva) samples for new assays. All data access policies and materials, including application forms, can be found at https://cls.ucl.ac.uk/data-access-training. A register of approved genetic projects is also maintained on the CLS data access page. The CLS DAC evaluates all requests following the principles and criteria outlined in the CLS Data Access Framework, ensuring that data are shared responsibly and securely and accessed only by worldwide bona fide researchers with due consideration to relevant ethical issues. Researchers should carefully review the CLS DAC data access guidelines and submit a completed application form to. Future access may—subject to their development— be via a Trusted Research Environment. For access to 1946c, see skylark.ucl.ac.uk.

## Ethics

Ethical approval was obtained in each study: 1946c (North Thames Multicentre Research Ethics Committee: reference 98/2/121 and 07/H1008/168), 1958c (South East Multi-centre Research Ethics Committee: ref 01/1/44), 1970c (South East Coast—Brighton & Sussex: ref 15/LO/1446), 1989c (East of England – Cambridge Central Research Ethics Committee: ref 22/EE/0052), 2001c (London-Central REC: 13/LO/1786).

## Acknowledgements

We thank colleagues in the Centre for Longitudinal Studies Research Data Management Team, Survey, Cohort Maintenance, Administrative, and Communications teams; as well as Next Steps PI Morag Henderson. We also thank colleagues at the University of Bristol for their work on biosample assay and storage.

## Contributions

Wrote the first draft: Gemma Shireby, David Bann, Tim Morris. All authors contributed to reviewing and revising the text. Liam Wright created Figure 1.

