## Supplementary Table 1 for "Data Resource Profile: Genomic Data in Multiple British Birth Cohorts (1946-2001)—Health, Social, and Environmental Data from Birth to Old Age"

**Supplementary Table 1. Sample sizes and those excluded at each stage of the quality control (QC) process.**

| **QC Step** | **1946 NSHD** | **1958 NCDS** | **1970 BCS** | **1989-90 Next Steps** | **Millennium cohort** |
| --- | --- | --- | --- | --- | --- |
| **Starting samples** | 2912 | 10829 | 5905 | 1683 | 21566 |
| **Read into GenomeStudio** | 2912 | 10829 | 5830 | 1681 | 21556 |
| **Individual-level exclusions (e.g. consent issues/ withdrawal)** | 0 | 67 | 0 | 0 | 348 |
| **>2% missing genotype rate per individual** | 102 | 204 | 136 | 57 | 667 |
| **mismatch sex** | 20 | 20 | 15 | 36 | 86 |
| **excess heterozygosity** | 12 | 61 | 46 | 18 | 78 |
| **mismatched samples based on KING** | NA | NA | NA | NA | 26 |
| **King related families** | NA | NA | NA | NA | 212 |
| **Duplicated samples excluded on merge** | NA | 4358 | NA | NA | NA |
| **King related individuals** | 1 | 21 | 35 | 5 | NA |
| **European** | 2731 | 6324 | 5423 | 1272 | 17,460 |
| **Total** | 2777 | 6396 | 5,598 | 1568 | 20,247 |

Note: values may differ in future (e.g., if participants withdraw consent or greater genotyping coverage is obtained). For updated sample sizes, please see: [https://cls-genetics.github.io/](https://cls-genetics.github.io/docs/intro.html)

NA values indicate QC steps not applicable to specific cohorts. Within-family KING analyses were exclusive to the Millennium cohort, while duplicate removal after chip merging applied only to the 1958 NCDS cohort. For the individual-level exclusions, there were multiple samples per individual removed across the multiple 1958 NCDS chips assayed separately.
